# Efficacy of a Filtered Far-UVC Handheld Disinfection Device on Reducing Microbial Bioburden of Hospital Surfaces

**DOI:** 10.1101/2022.11.07.22282040

**Authors:** Thanuri Navarathna, Chetan Jinadatha, Brandon A. Corona, John David Coppin, Hosoon Choi, Morgan R. Bennett, Gautam S. Ghamande, Marjory D. Williams, Robin E. Keene, Piyali Chatterjee

## Abstract

**Objectives:** The Filtered Far-UVC (FFUV) handheld disinfection device is a small portable device that emits far UVC at 222nm. The objective of this study was to evaluate the device’s ability to kill microbial pathogens on hospital surfaces and compare it to manual disinfection using germicidal sodium hypochlorite wipes.

**Methods:** A total of 344 observations (4 observations from 86 objects’ surfaces) were sampled with 2 paired samples per surface: a pre- and a post-sodium hypochlorite, and a pre- and a post-FFUV samples. The results were analyzed via a Bayesian multilevel negative binomial regression model. Additionally, the bacterial flora recovered were identified using mass spectrometry.

**Results:** The estimated mean colony counts for the sodium hypochlorite control and treatment groups were 20.5 (11.7 – 36.0) and 0.1 (0.0 – 0.2) colony forming units (CFUs) respectively. The FFUV control and treatment groups had mean colony counts of 22.2 (12.5 – 40.1) and 4.1 (2.3 – 7.2) CFUs. The sodium hypochlorite samples had an estimated 99.4% (99.0% – 99.7%) reduction in colony counts, while those from the FFUV group had an 81.4% (76.2% − 85.7%) reduction.

**Conclusions:** Our study demonstrated that FFUV handheld device effectively reduced microbial bioburden on surfaces in the healthcare setting. Several healthcare-associated infections (HAIs) causing pathogens (gram positive and negative bacteria) were retrieved from the pre-clean surfaces. The major benefit of FFUV is likely seen when manual disinfection is not possible or when supplementing cleaners or disinfectants with the low-level disinfection properties.

## Introduction

Healthcare equipment and surfaces can harbor harmful pathogens that can cause healthcare-associated infections because of constant interaction between hands of healthcare workers, surfaces, and equipment.^1^ Many handheld ultraviolet devices are effectively used in the healthcare setting to disinfect equipment and surfaces.^2,3,4,5^ However, certain UV-C wands used for disinfecting surfaces may expose both the end user and those nearby to unsafe levels of UV-C radiation that can harm the skin and eyes within a few seconds of exposure.^6,3^ Thus, there is a need for devices that are both safe for users yet effective in disinfecting surfaces or equipment.

The Filtered Far-UV-C (FFUV) handheld disinfection device produces UV light at a wavelength of 222nm that is germicidal with short contact time but without the drawbacks of 254nm UV-C devices. At 222nm wavelength, FFUV has been shown in numerous studies to be potentially germicidal and safe for humans because of the very low penetration depth of FFUV to human skin and eyes at 222nm.^7, 8, 9, 10, 11, 12, 13^ *In vitro* evaluations indicate 222nm wavelengths are effective in deactivating a multitude of pathogens (both growth/stationary phase or spores) including methicillin-resistant *Staphylococcus aureus* (MRSA), *Bacillus cereus, Bacillus subtilis, Bacillus thuringiensis, Clostridioides difficile*, and Herpesvirus.^14,15^ However, no data are available that attempt to determine if the FFUV handheld disinfection device will be effective or practical for use on different types of surfaces in healthcare settings and compared to standard manual cleaning to eliminate bacterial pathogens.

In this study, we evaluated the ability of a FFUV handheld disinfection device to reduce bacteria commonly found on surfaces in the healthcare setting and compared it to using a germicidal sodium hypochlorite disinfectant wipe.

## Methods

The study was conducted in an acute care facility (Central Texas Veterans Health Care System) in Temple, TX with approvals from the Research and Development committee.

### The FFUV handheld disinfection device

The FFUV handheld disinfection device used for the study was developed by Freestyle Partners, LLC, and its affiliate, FSP Innovations, LLC which included the Ushio Care222 lamp technology (Figure 1A-D). The unit contains four highly efficient 222nm excimer lamps. The excimer lamp, 12W B1 module, contains a chamber filled with a noble gas mixture (Kr-Cl gas) that does not use inner electrodes or contain mercury. When high voltage is applied across the outside of the glass, it “excites” the gaseous mixture inside emitting UV light. A patented optical filter then eliminates the harmful longer wavelengths of more than 230nm. The device emits ~3.6 mW/cm^2^ at a 1-inch distance from the intended surface, which is within the current 2022 exposure limits recommended by the American Conference of Governmental Industrial Hygienists (ACGIH®) and American National Standards Institute (ANSI) /IES (Illuminating Engineering Society) RP 27.1-22, when used as intended. A green light (Figure 1D) on the handheld device indicates when the intended operating distance of 1” inch (2.54 cm) from the target surface is reached. Prior to each use, both the dosage and the wavelength of emission of the FFUV handheld disinfection device were validated (Figure 1).

**Figure 1.**
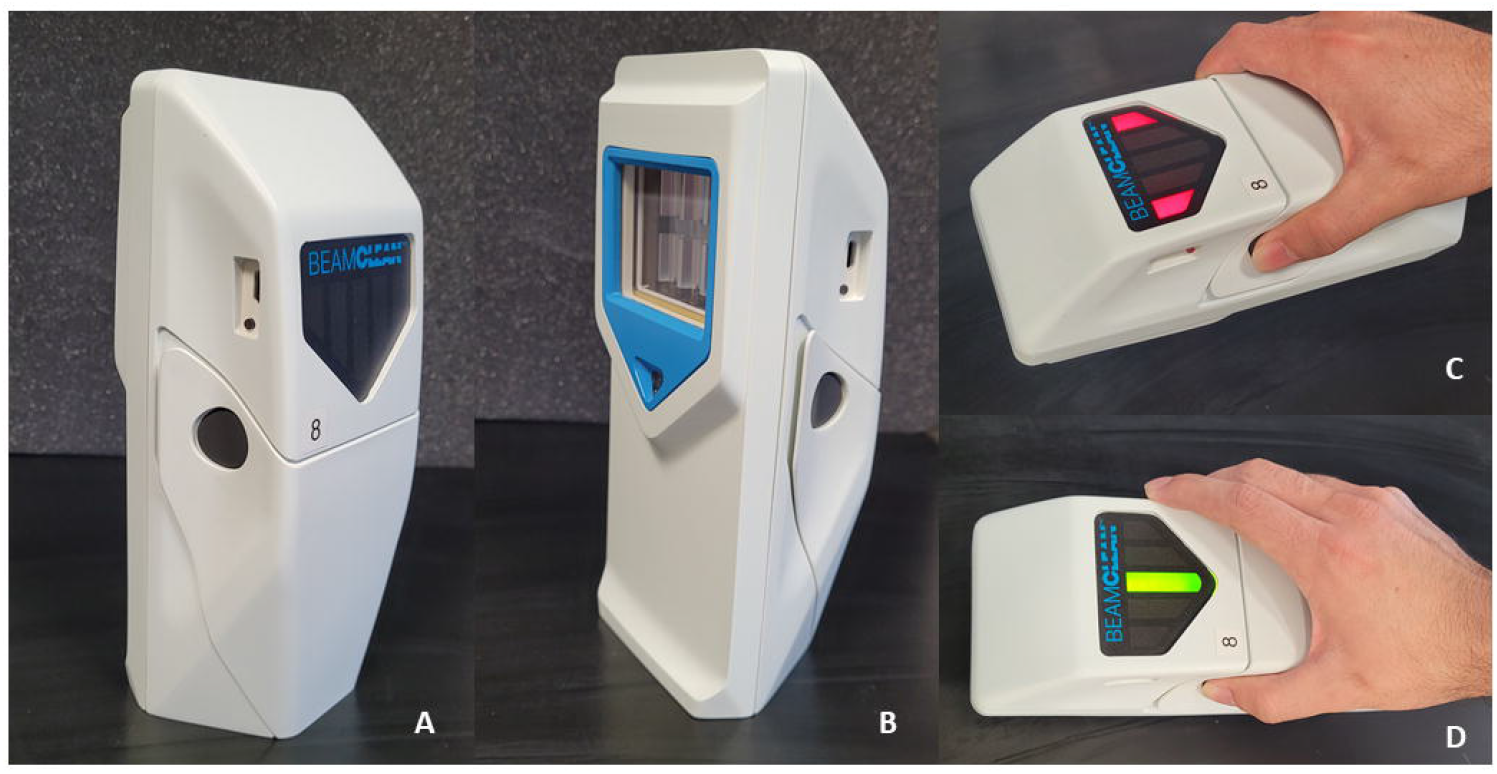
The portable battery-operated FFUV handheld disinfection device A) side view showing the “on” button (round black) on the device that needs to be held down during operation B) side view of the device showing the glass surface area in the bottom that emanates the UV-C light C) The “red indicator light” suggests the correct distance for the surface to be disinfected has been not yet achieved. D) The “green indicator light” indicates the correct distance has been reached.

### Sample Collection

All the study samples were collected between June-July 2022 in various inpatient medical surgical units. The study had 2 arms: The FFUV arm and sodium hypochlorite based disinfectant wipe arm. Each arm had matched pre-post samples from the same surface. Several high-touch surfaces: bedrails, computer keyboards at nurse’s stations, simulation manikins, breakroom tables, and workstations-on-wheels (WOWs) were selected for sampling due to high frequency of contact.^1^ For each surface a corresponding pre-post sample was collected for both the FFUV (30 seconds exposure) and disinfectant arm (4 mins dwell time), for a total of 4 samples per surface. A total of 86 high-touch surfaces across study units were sampled. Due to known impenetrability of UV when organic material is present, grossly soiled surfaces were avoided. Rodac tryptic soy agar (Hardy Diagnostics, Santa Maria, CA, USA) contact plates (25cm^2^ surface area) were used for sampling as described previously.^16^ Plates were subsequently incubated in aerobic conditions at 35°C+/-2°C for 24 hours, and colony counts were enumerated. One representative sample from each morphology type was sub-cultured on a blood agar plate (TSA with sheep blood, Thermofisher Scientific) and speciated using Matrix-assisted laser desorption ionization-time of flight mass spectrometry (MALDI-TOF MS, Biomerieux). Colonies identified as *Staphylococcus aureus* were further grown on Columbia CNA agar w/5% sheep blood plates (Remel) and further characterized as methicillin-resistant *Staphylococcus aureus* (MRSA) by using two different kits, PBP2a SA culture colony test (Abbott) and Sure-Vue color staph ID latex test kit (Fisher Healthcare), following manufacturer’s instructions.

Statistical analysis was conducted using a Bayesian multilevel negative binomial regression model, since the outcome consisted of over-dispersed counts. The model was run in the ‘brms’ package version 2.17.0 in R version 4.1.3. All plots were created in the ‘ggplot2’ package.

## Results

### Efficacy of Disinfection

A total of 344 samples were available for final analysis. The colony counts for the controls in disinfectant wipe arm ranged from zero colonies to 155 colonies with a median of 20. Similarly, the controls in the FFUV arm ranged from zero to 140 colonies with a median of 16.5. Post disinfection colony counts ranged from 0-2 for disinfectant wipe arm and 0-76 for the post-FFUV arm. The estimated mean colony counts for the pre- and post-sodium hypochlorite disinfectant arm was 20.5 (95% uncertainty interval: 11.7 to 36.0) and 0.1 (0.0 – 0.2) colony forming units (CFUs), respectively. The Bayesian 95% uncertainty interval, for example 11.7-36.0, can be interpreted as a 95% chance that the mean counts fall in that interval, conditional on our data and model. Similarly, for the FFUV arm the mean colony counts were 22.2 (12.5 – 40.1) and 4.1 (2.3 – 7.2) CFUs, respectively (**Figure 2**). Samples in the sodium hypochlorite group had a 99.4% (99.0% – 99.7%) reduction in colony counts, while the samples in the FFUV group had an 81.4% (76.2% − 85.7%) reduction in colony counts. Both the interventions (manual disinfection and FFUV) had a substantial effect in reducing bioburden (**Figure 2**) but the manual disinfection arm was better than the FFUV arm as expected. We recovered several pathogenic gram-positive and gram-negative aerobes from pre-clean surfaces. Gram-positive bacteria such as *Staphylococcus, Bacillus*, and *Micrococcus* and Gram-negative bacteria such as *Acinetobacter, Citrobacter, Pseudomonas, Klebsiella, and Escherichia* were among others that were found on the surfaces. Many of these pathogenic bacteria were found on all types of high-touch surfaces that were sampled (**Figure 3**).

**Figure 2.**
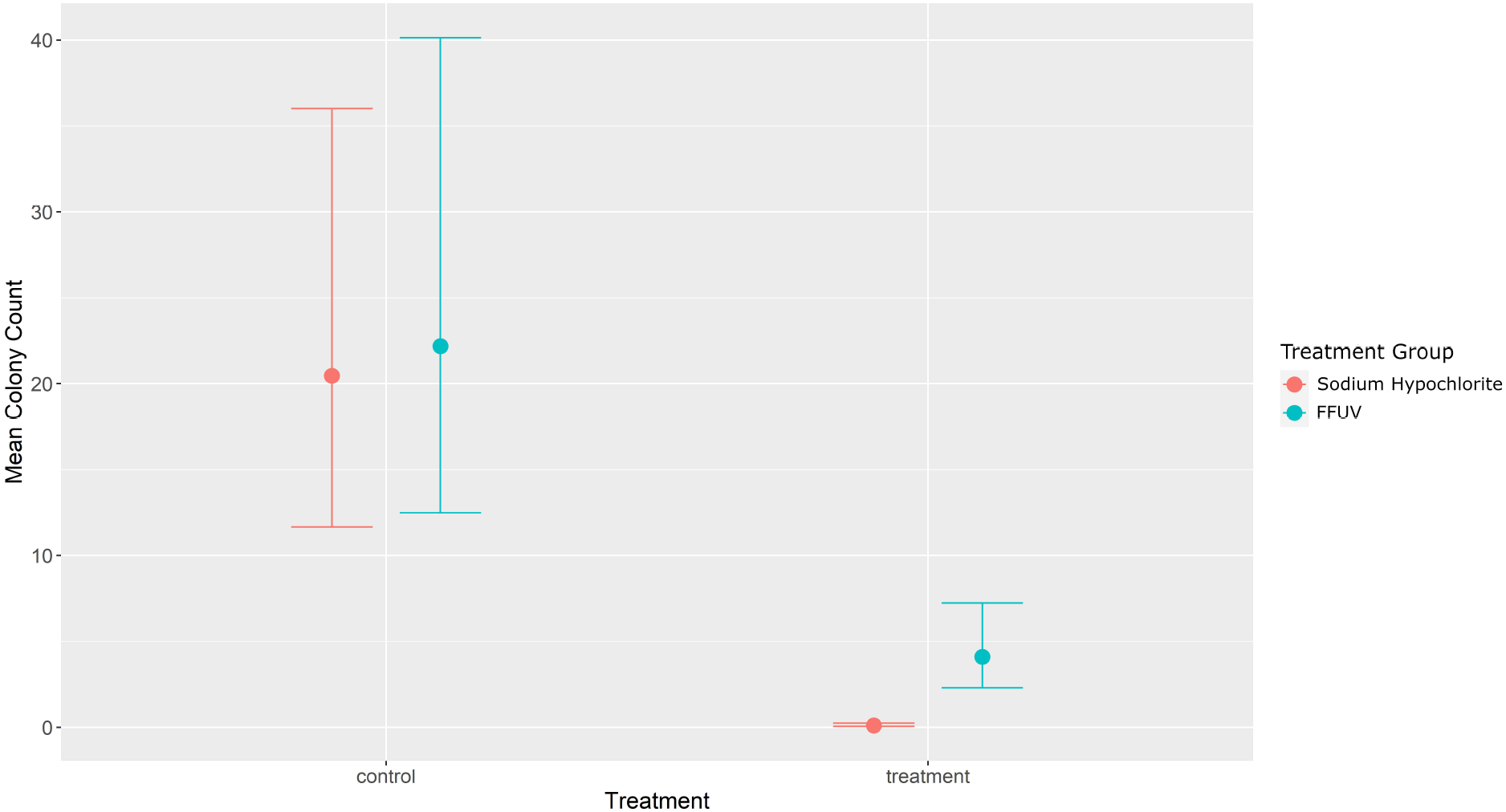
Model estimated mean (points) and 95% Uncertainty Intervals (whiskers) for the sodium hypochlorite (red) and UV (green) control (left) and treatment (right) groups.

**Figure 3.**
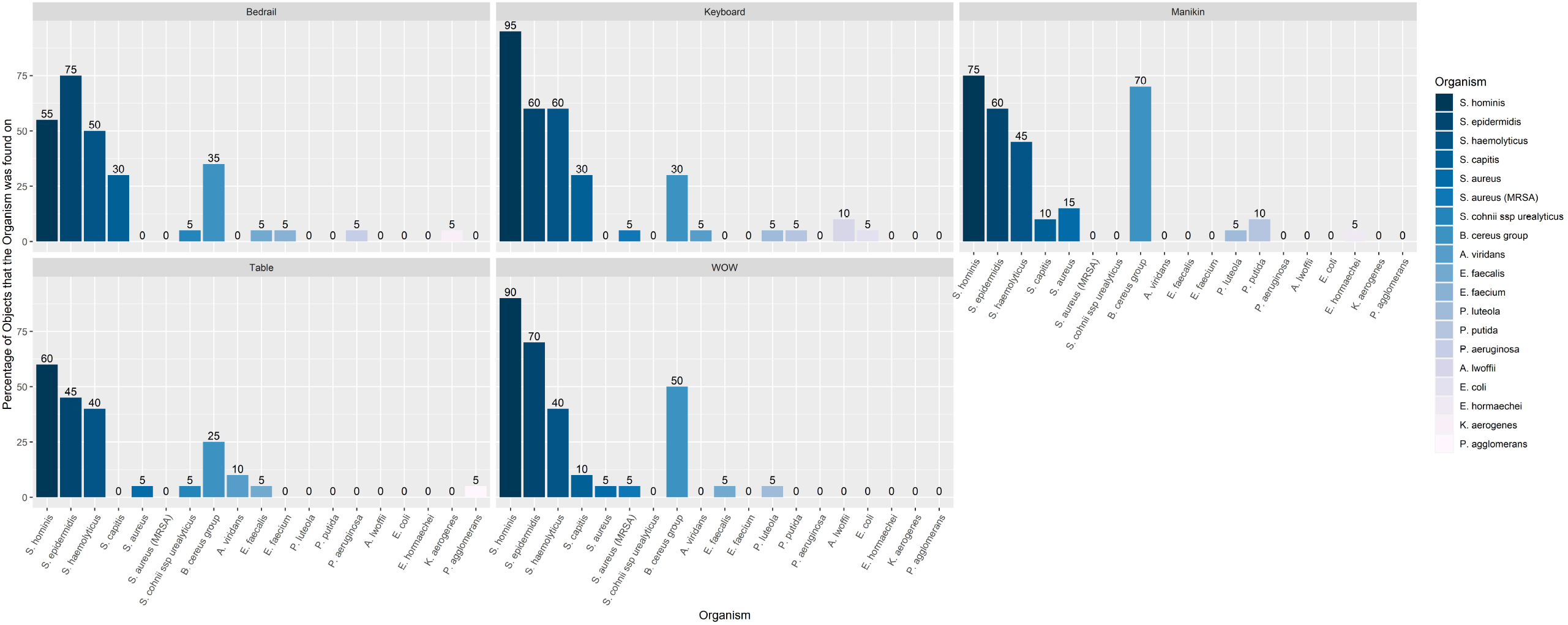
Organisms recovered from surfaces of the 5 objects grouped by gram stain and ordered by percentage in descending order within genus.

## Discussion

The FFUV handheld disinfection device used in this study demonstrated the ability to reduce microbial bioburden on surfaces in a real-world healthcare environment. The reduction in colonies post-FFUV disinfection was substantial and comparable to other handheld UV devices tested previously.^2,4,3,5^ The effect of UV is dependent on dose and distance. Unlike other handheld UV devices, the FFUV handheld disinfection device detects the exact distance from the surface with an indicator for application, resulting in a consistent dose during use while providing guidance to the end users for optimal results.^2^ Many handheld UV devices that are currently commercially available have not undergone rigorous peer reviewed scientific testing in either controlled or real-world testing in a healthcare setting. Additionally, there have been several concerns regarding the safety of handheld UV devices and many of them have centered around inadvertent exposure of UV to users. The FDA issued a safety warning about handheld wands, alerting users that UV-C radiation may harm eyes and skin within a few seconds of exposure.^3,6^ The FFUV dose administered for this study is known to be safe for humans.^7,8^

In our current study, the sodium hypochlorite-based wipe disinfectant was superior to FFUV disinfection alone in its ability to disinfect. The advantages of a wipe-based disinfectant include abilities to remove both organic material (dirt and debris) and organisms with mechanical action as well as chemical disinfectant properties (oxidation). Several studies have shown that manual disinfection alone is not always adequate for high-touch surfaces especially with low-level disinfectants, such as quaternary ammonium compounds or alcohol-based disinfectants, due to issues with resistance to disinfectants or user errors.^17,16,18^ The use of low-level disinfection cleaners followed by UV-C use provided a greater benefit than use of high disinfection property cleaners such as 10% sodium hypochlorite or hydrogen peroxide-based disinfectants followed by UV-C.^16^ Our previous UV studies for whole room disinfection showed that using UV-C after manual disinfection decreased residual bacterial counts more than manual disinfection alone,^16^ and the overall contribution of UV-C to the disinfection process was dependent on the type of cleaning disinfectant that preceded the UV-C use. One disadvantage of using sodium hypochlorite wipes is that they cannot be used on certain sensitive equipment due to non-compatibility or the risk of voiding manufacturer warranty, which may force end users to use potentially inadequate or ineffective disinfectant wipes.^19^ In those instances, FFUV handheld disinfection device has the potential to act as a method of disinfection for these healthcare surfaces, without added safety concerns for users or damage for equipment and surfaces.

Although we used FFUV alone in our experiments (no manual disinfection before FFUV), we still recommend that FFUV be preceded by any sort of manual cleaning or disinfection using wet wipes that are compatible as recommended by the equipment manufacturer before using FFUV. The UV-C disinfection (FFUV or otherwise) does not have the advantage of mechanical removal of organisms. The combined effect of manual disinfection and FFUV should achieve better results than manual disinfection or FFUV alone. But, if any wet wipe down of equipment is specifically prohibited by manufacturer, we recommend using FFUV independently because of its potential to reduce bioburden even without manual wipe down. Grossly contaminated areas with visible dirt will need wipe down regardless of FFUV use, as UV is ineffective in penetrating organic debris.^2^

Microbial bioburden including the pathogenic organisms recovered in our hospital environment may be largely a product of the frequency of touches, thus increased frequency of cleaner and disinfectant use may have more impact on the bioburden at any given time.^1^ Whether more frequent use of a UV device may be more convenient for end users than use of chemical disinfectants that have certain limitations needs to be further evaluated in future larger studies.^19^

Our study has some limitations. The contact plates used for our study may not have been able to capture all bacteria such as fastidious or anaerobes. It is also difficult to sample uneven surfaces using contact plates. The study data was collected by trained research personnel (not end users) to have consistency of device use and disinfectant wipes, which usually yields superior results compared to regular users.^20^ We did not study the impact of FFUV on healthcare-associated infection rates. Additional large-scale studies are also needed to determine if the FFUV handheld disinfection device can reduce healthcare-associated infections.

## Conclusions

The FFUV handheld disinfection device can effectively reduce bioburden on surfaces. The largest effect is likely seen when manual disinfection is not possible or when supplementing cleaners or disinfectants with the lowest disinfection properties. It is also a safer alternative to other handheld UV-C devices due to filtration of UV-C known to be harmful to end users.

## Data Availability

All data produced in the present study are available upon reasonable request to the authors.

## Acknowledgements

The authors appreciate the support from all the nursing unit managers and staff for allowing research personnel to conduct study activities planned in this manuscript. Special thanks to Kristy N. Causey and the Center for Innovation and Learning and Gracia M. Boseman, Infection Prevention and Control for their support of our study. The authors thank Freestyle Partners, LLC, and its affiliate, FSP Innovations, LLC for providing the devices and for use of the calibration verification device.

## Author contributions

TN, BAC collected samples; TN, BAC, HC, and GG processed specimens in lab; JDC provided statistical analysis and generated figures; all authors participated in the study design, data analysis, and editing of the manuscript. PC wrote the manuscript and provided overall guidance for the project. All authors have read and approved the submitted manuscript.

